# Impact of Significant Hemoglobin Drop without Bleeding in Patients Undergoing Transcatheter Aortic Valve Replacement

**DOI:** 10.1101/2023.08.11.23294000

**Authors:** Pavan Reddy, Ilan Merdler, Cheng Zhang, Matteo Cellamare, Itsik Ben-Dor, Lowell F. Satler, Toby Rogers, Hector M. Garcia-Garcia, Ron Waksman

**Author notes:** **Address for Correspondence** Ron Waksman, MD MedStar Washington Hospital Center 110 Irving St. NW, Suite 4B-1 Washington, DC 20010.

## Abstract

**Background:** Hemoglobin (Hgb) drop without bleeding is common among patients undergoing transcatheter aortic valve replacement (TAVR); however, the clinical implications of significant Hgb drop have not been fully evaluated. We sought to describe clinical phenotype and outcomes of patients with significant Hgb drop without bleeding.

**Methods:** Consecutive patients undergoing TAVR at our institution from 2011-2021 were retrospectively reviewed. Three groups were assessed: No Hgb Drop and No Bleed (NoD-NoB – reference group), Hgb Drop with Bleed (D-wB), and Hgb Drop and No Bleed (D-NoB). Hgb drop was defined as ≥3 g/dL decrease from pre-to post-TAVR. Outcomes of interest were in-hospital death and 1-year all-cause mortality.

**Results:** A total of 1,851 cases with complete Hgb data were included: NoD-NoB: n=1,579 (85.3%); NoD-NoB: n=49 (2.6%); D-wB n=223 (12.6%). Compared to NoD-NoB, D-NoB were older (81.1 vs. 78.9 years) with higher pre-procedure Hgb (12.9 vs. 11.7 g/dL). In-hospital death rate was higher among patients with D-NoB vs. NoD-NoB (4.5% vs. 0.8%, p<0.001) and similar to D-wB (4.5% vs. 4.1%, p=0.999). Predictors of in-hospital death were D-NoB (odds ratio [OR]: 3.45; 95% confidence interval [CI]: 1.32-8.69) and transfusion (OR: 10.6; 95% CI 4.25-28.2). Landmark survival analysis found that D-NoB experienced one-year mortality rate comparable to NoD-NoB, while D-wB had higher mid-term mortality (HR: 3.2; 95% CI: 1.83-5.73), and transfusion continued to impact mortality (HR: 2.5; 95% CI 1.79-3.63).

**Conclusion:** Hemoglobin drop without bleeding is common among patients undergoing TAVR and may represent a higher risk of peri-procedural death. Blood transfusion increases short-and mid-term mortality risk in patients with and without bleeding, supporting a restrictive transfusion strategy.

**What Is Known; What the Study Adds:** **What is Known**

- Hemoglobin drop without overt bleeding may be seen after transcatheter aortic valve replacement (TAVR); however, its incidence and impact on clinical outcomes is unknown.

**What the Study Adds**

- Hemoglobin drop without a source of bleeding is not uncommon among patients undergoing TAVR.
- Patients more likely to have hemoglobin drop without bleeding include those with lower body surface area, older age, and higher pre-procedural hemoglobin level.
- Compared to patients without bleeding or hemoglobin drop, patients with hemoglobin drop without bleeding had higher inpatient mortality.
- Blood transfusion increased short-and long-term mortality in patients with or without bleeding.

**Visual Abstract:** 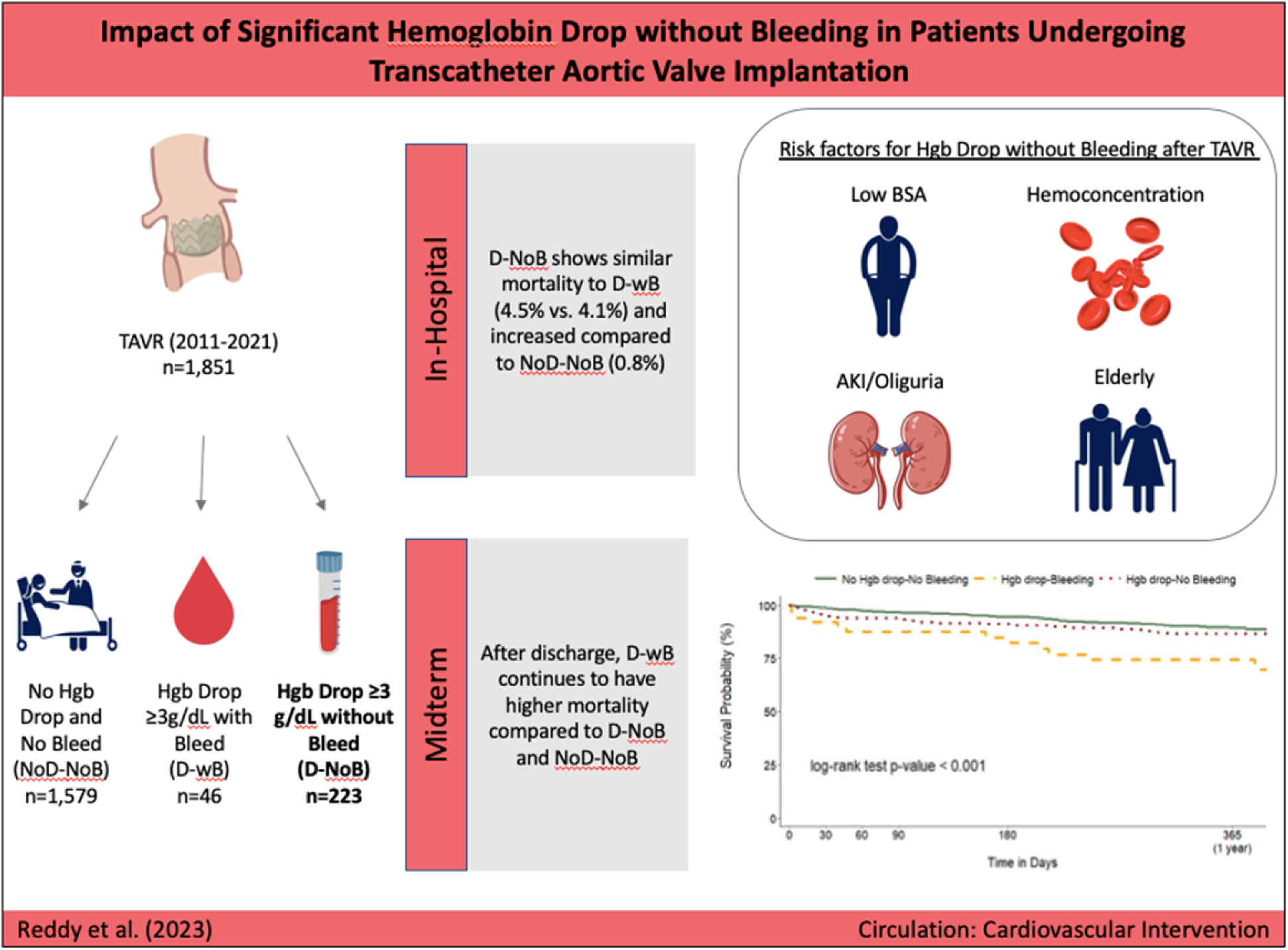

## Introduction

Transcatheter aortic valve replacement (TAVR) is commonly performed in patients at high bleeding risk secondary to advanced age and multiple comorbidities^1^. Bleeding events are well known to adversely affect the clinical course of patients after TAVR; their effect may be further risk-stratified by the change in hemoglobin (Hgb), i.e., the degree of blood loss^2^. However, Hgb drop without overt bleeding is common among hospitalized patients, often occurring due to a combination of poor bone marrow production with repeated phlebotomy or minor access-site bleeding, or from a dilutional-effect intravenous saline^3, 4^. In the setting of recent TAVR, Hgb drop can raise particular concern, causing reactionary orders that can adversely affect overall outcomes such as blood transfusion or changes to medication regimens^5^. Current data are limited regarding the effect of Hgb drop without bleeding on clinical outcomes after TAVR.

This study aimed to: 1) identify which patients undergoing TAVR are more likely to have Hgb drop without bleeding, and 2) determine the impact of Hgb drop without bleed on peri-procedural and mid-term clinical outcomes.

## Methods

### Study Design

This study was approved by the institutional review board at MedStar Washington Hospital Center. We retrospectively identified consecutive patients who underwent TAVR between 2011 and 2021 using institutional data entered into the Transcatheter Valve Therapy (TVT) Registry. Three groups were defined: No Hgb drop and No Bleed (NoD-NoB – reference group), Hgb drop with bleed (D-wB), and Hgb drop and no bleed (D-NoB). Bleeding was defined by the presence of a recorded bleeding event from adjudicated data in the TVT Registry combined with an Hgb drop ≥3 g/dL from pre-procedure Hgb to the lowest Hgb post-procedure. The D-NoB group was defined by Hgb drop ≥3 g/dL without any reported bleeding event. An Hgb drop of ≥3 g/dL was used as the benchmark in this study in accordance with Valve Academic Research Consortium (VARC) 3 criteria for “major bleeding,” because it marks the threshold for increased risk of adverse clinical outcomes.^2, 6^ Patients noted to have had bleeding but without evidence of Hgb drop were excluded from the analysis to facilitate isolating the effect of Hgb drop with vs. without bleeding. Qualifying bleeding events in the TVT Registry include gastrointestinal bleed, access-site bleed, hematoma at access site, and retroperitoneal bleed. Records were also excluded if Hgb data were incomplete or if deemed erroneous (Hgb drop >7 or a pre-procedure Hgb of <5 g/dL or >18 g/dL). The data that support the findings of this study are available from the corresponding author upon reasonable request.

### Clinical Endpoints

The main outcomes of interest were in-hospital and 1-year rates of all-cause death. Secondary outcomes included hospital course events (acute kidney injury [AKI] and blood transfusion) and bleeding events at follow up. AKI was determined from pre-and post-procedure creatinine and defined according to VARC 3 guidelines for stage 1 AKI. Follow-up visits occurred at 1 month and 1 year post-procedure, either face to face or by telephone. Adverse clinical events were ascertained at that time by the treating physician.

### Statistical Analysis

Continuous variables are presented as mean ± standard deviation and categorical variables as proportions. Student t test and χ^2^ test were used to evaluate the statistical significance between continuous and categorical variables, respectively. Multivariate logistic regression was used to assess the effect of Hgb drop with or without bleeding on in-hospital death within the context of other clinically relevant variables. Due to the small number of events, a limited number of predetermined covariates (transfusion and pre-procedure anemia) were included in the model. A second model with a greater number of variables is presented in Table S1). Odds ratios were calculated with a confidence interval (CI) of 95%.

Kaplan-Meier curves were used to estimate event rates during follow-up. Given the observed high rate of in-hospital death in the D-wB and D-NoB groups, we performed Kaplan-Meier landmark analysis, designating the day of discharge as day 0, to isolate the effect of Hgb drop and bleeding on mid-term outcomes. Unadjusted 1-year mortality and bleeding were obtained from Cox proportional hazards models with the three cohorts as the only covariate. Further survival analyses were performed with the use of multivariate Cox proportional hazards analyses, adjusted for clinically relevant variables. The proportional hazard assumption was verified in all Cox analyses. The results were displayed as hazard ratio (HR) with its 95% CI. All analyses were considered significant for P<0.05 (2-sided). All analyses were performed with R (v 4.1.3) and SAS 9.2 (SAS Institute, Cary, NC).

## Results

Query of the TVT Registry identified 2,153 TAVR cases completed at our institution from 2011 to 2021. Of these, 69 cases with reported bleeding events were excluded because they did not demonstrate an Hgb drop of ≥3 g/dL; 233 patients were excluded for incomplete Hgb data. Thus, 1,851 cases with complete Hgb data were included in the analysis: 1,579 (85.3%) patients had NoD-NoB; 49 (2.6%) patients had D-wB; and 223 (12.6%) patients had D-NoB. *Baseline Characteristics (Table 1)*

**Table 1:**
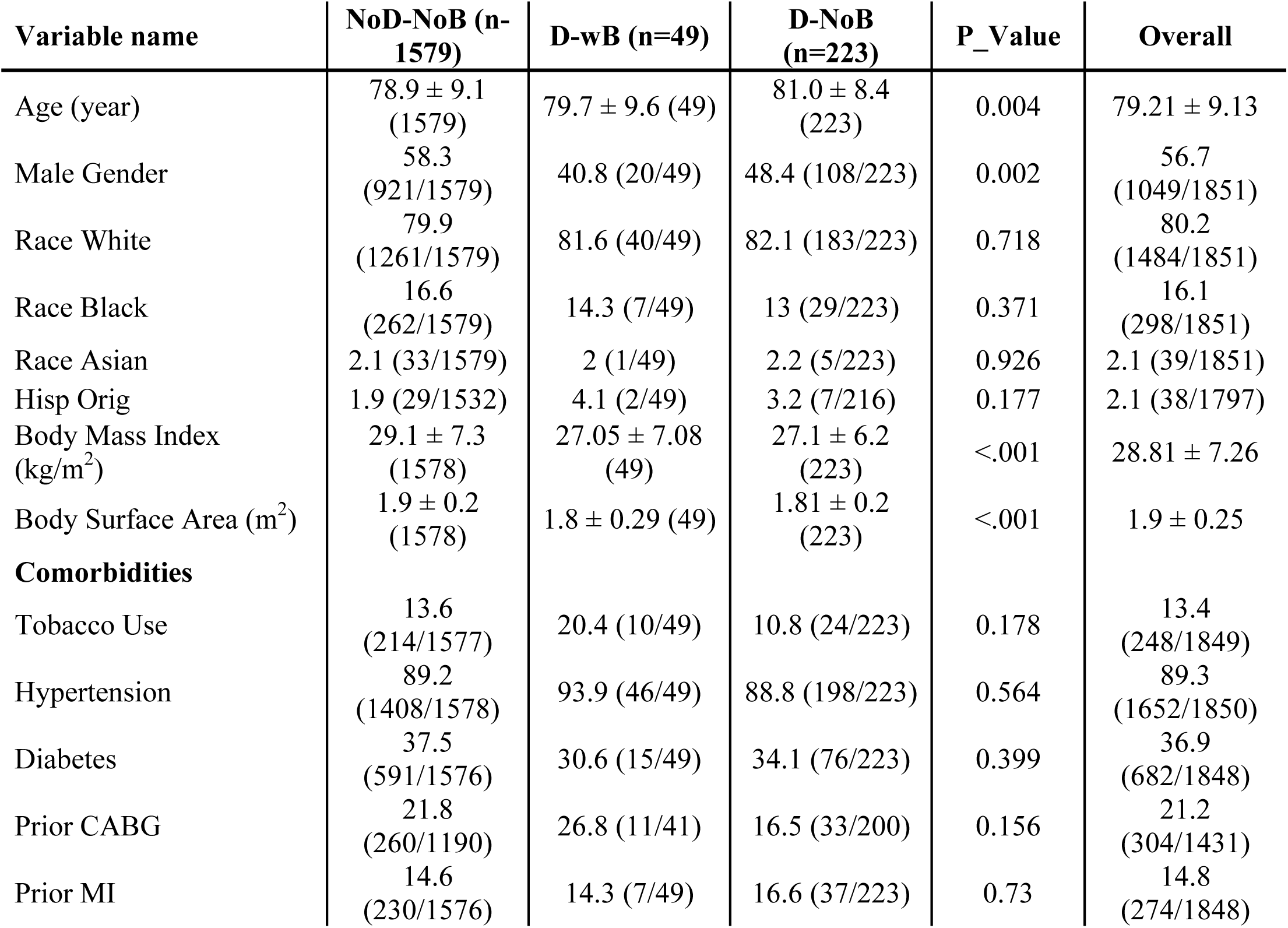
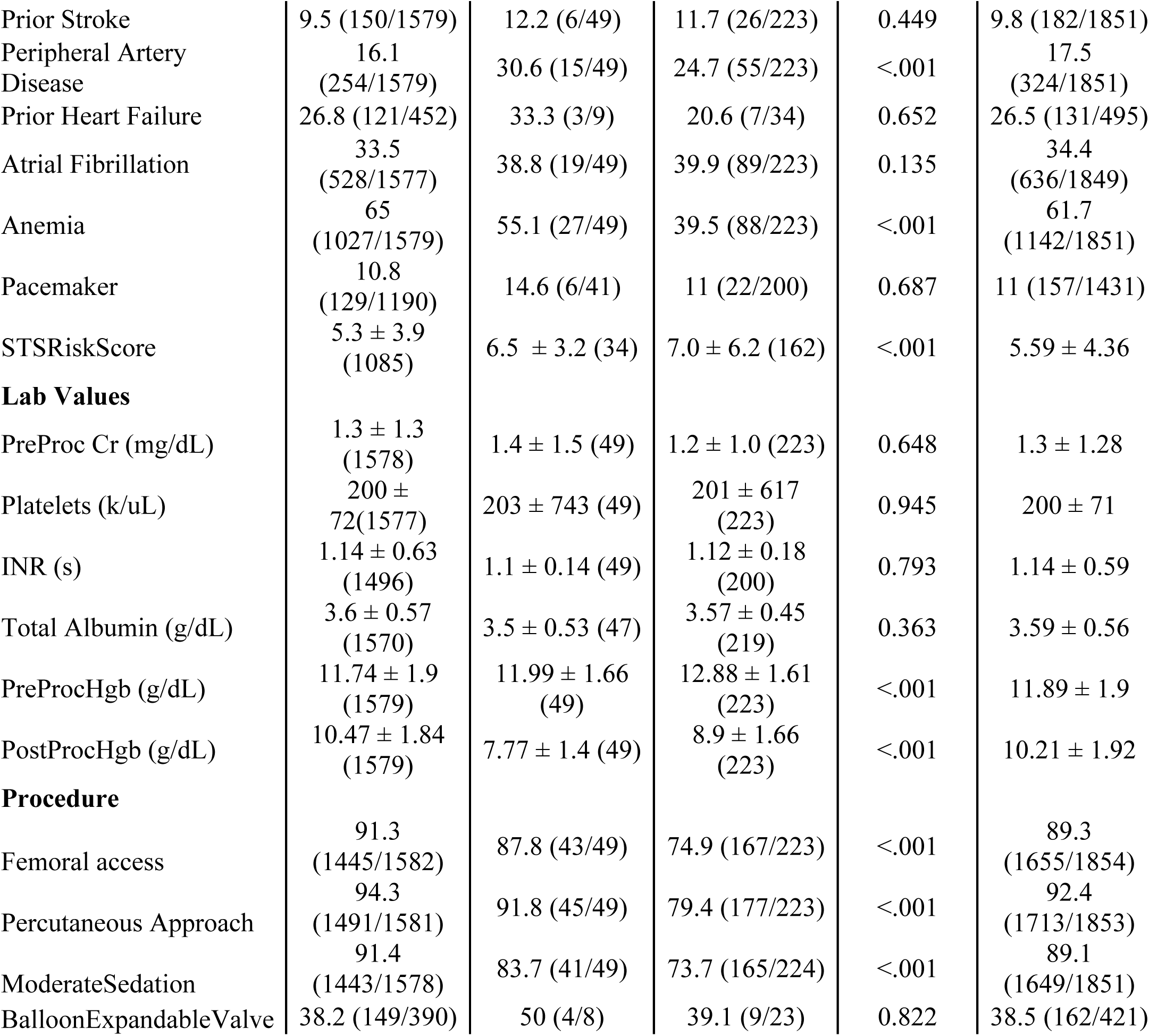
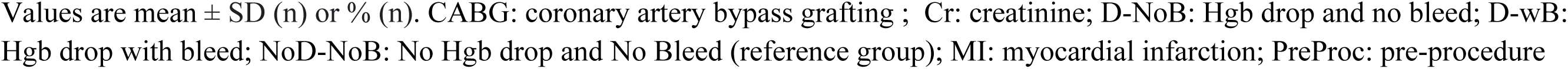
Baseline Characteristics.

| Variable name | NoD-NoB (n-1579) | D-wB (n=49) | D-NoB (n=223) | P_Value | Overall |
| --- | --- | --- | --- | --- | --- |
| Age (year) | 78.9 ± 9.1<br>(1579) | 79.7 ± 9.6 (49) | 81.0 ± 8.4<br>(223) | 0.004 | 79.21 ± 9.13 |
| Male Gender | 58.3<br>(921/1579) | 40.8 (20/49) | 48.4 (108/223) | 0.002 | 56.7<br>(1049/1851) |
| Race White | 79.9<br>(1261/1579) | 81.6 (40/49) | 82.1 (183/223) | 0.718 | 80.2<br>(1484/1851) |
| Race Black | 16.6<br>(262/1579) | 14.3 (7/49) | 13 (29/223) | 0.371 | 16.1<br>(298/1851) |
| Race Asian | 2.1 (33/1579) | 2 (1/49) | 2.2 (5/223) | 0.926 | 2.1 (39/1851) |
| Hisp Orig | 1.9 (29/1532) | 4.1 (2/49) | 3.2 (7/216) | 0.177 | 2.1 (38/1797) |
| Body Mass Index<br>(kg/m<sup>2</sup>) | 29.1 ± 7.3<br>(1578) | 27.05 ± 7.08<br>(49) | 27.1 ± 6.2<br>(223) | <.001 | 28.81 ± 7.26 |
| Body Surface Area (m<sup>2</sup>) | 1.9 ± 0.2<br>(1578) | 1.8 ± 0.29 (49) | 1.81 ± 0.2<br>(223) | <.001 | 1.9 ± 0.25 |
| **Comorbidities** |  |  |  |  |  |
| Tobacco Use | 13.6<br>(214/1577) | 20.4 (10/49) | 10.8 (24/223) | 0.178 | 13.4<br>(248/1849) |
| Hypertension | 89.2<br>(1408/1578) | 93.9 (46/49) | 88.8 (198/223) | 0.564 | 89.3<br>(1652/1850) |
| Diabetes | 37.5<br>(591/1576) | 30.6 (15/49) | 34.1 (76/223) | 0.399 | 36.9<br>(682/1848) |
| Prior CABG | 21.8<br>(260/1190) | 26.8 (11/41) | 16.5 (33/200) | 0.156 | 21.2<br>(304/1431) |
| Prior MI | 14.6<br>(230/1576) | 14.3 (7/49) | 16.6 (37/223) | 0.73 | 14.8<br>(274/1848) |
| Prior Stroke | 9.5 (150/1579) | 12.2 (6/49) | 11.7 (26/223) | 0.449 | 9.8 (182/1851) |
| Peripheral Artery Disease | 16.1 (254/1579) | 30.6 (15/49) | 24.7 (55/223) | <.001 | 17.5 (324/1851) |
| Prior Heart Failure | 26.8 (121/452) | 33.3 (3/9) | 20.6 (7/34) | 0.652 | 26.5 (131/495) |
| Atrial Fibrillation | 33.5 (528/1577) | 38.8 (19/49) | 39.9 (89/223) | 0.135 | 34.4 (636/1849) |
| Anemia | 65 (1027/1579) | 55.1 (27/49) | 39.5 (88/223) | <.001 | 61.7 (1142/1851) |
| Pacemaker | 10.8 (129/1190) | 14.6 (6/41) | 11 (22/200) | 0.687 | 11 (157/1431) |
| STSRiskScore | 5.3 ± 3.9 (1085) | 6.5 ± 3.2 (34) | 7.0 ± 6.2 (162) | <.001 | 5.59 ± 4.36 |
| **Lab Values** |  |  |  |  |  |
| PreProc Cr (mg/dL) | 1.3 ± 1.3 (1578) | 1.4 ± 1.5 (49) | 1.2 ± 1.0 (223) | 0.648 | 1.3 ± 1.28 |
| Platelets (k/uL) | 200 ± 72 (1577) | 203 ± 743 (49) | 201 ± 617 (223) | 0.945 | 200 ± 71 |
| INR (s) | 1.14 ± 0.63 (1496) | 1.1 ± 0.14 (49) | 1.12 ± 0.18 (200) | 0.793 | 1.14 ± 0.59 |
| Total Albumin (g/dL) | 3.6 ± 0.57 (1570) | 3.5 ± 0.53 (47) | 3.57 ± 0.45 (219) | 0.363 | 3.59 ± 0.56 |
| PreProcHgb (g/dL) | 11.74 ± 1.9 (1579) | 11.99 ± 1.66 (49) | 12.88 ± 1.61 (223) | <.001 | 11.89 ± 1.9 |
| PostProcHgb (g/dL) | 10.47 ± 1.84 (1579) | 7.77 ± 1.4 (49) | 8.9 ± 1.66 (223) | <.001 | 10.21 ± 1.92 |
| **Procedure** |  |  |  |  |  |
| Femoral access | 91.3 (1445/1582) | 87.8 (43/49) | 74.9 (167/223) | <.001 | 89.3 (1655/1854) |
| Percutaneous Approach | 94.3 (1491/1581) | 91.8 (45/49) | 79.4 (177/223) | <.001 | 92.4 (1713/1853) |
| ModerateSedation | 91.4 (1443/1578) | 83.7 (41/49) | 73.7 (165/224) | <.001 | 89.1 (1649/1851) |
| BalloonExpandableValve | 38.2 (149/390) | 50 (4/8) | 39.1 (9/23) | 0.822 | 38.5 (162/421) |

Values are mean ± SD (n) or % (n). CABG: coronary artery bypass grafting ; Cr: creatinine; D-NoB: Hgb drop and no bleed; D-wB: Hgb drop with bleed; NoD-NoB: No Hgb drop and No Bleed (reference group); MI: myocardial infarction; PreProc: pre-procedure

All three groups were similar in prevalence of comorbidities, with the exception of peripheral artery disease, which was higher in both the D-wB and D-NoB groups compared to the reference group (NoD-NoB). Platelet count, international normalized ratio, and pre-procedure creatinine levels were also similar among the groups. The following differences were observed between the D-NoB group and the reference group: D-NoB were older (81.1 years vs. 78.9 years) with lower body surface area (BSA) (1.81 m^2^ vs. 1.92 m^2^), higher pre-procedure Hgb (12.9 g/dL vs. 11.7 g/dL), and higher Society of Thoracic Surgeons Predicted Risk of Mortality (STS-PROM) score (7.03 vs. 5.35, p<0.001 for all). While the D-NoB and D-wB groups were largely similar, the D-NoB group had higher pre-procedure Hgb (12.9 g/dL vs. 12.0 g/dL, p=0.002).

### Clinical Outcomes

In-hospital death occurred more frequently in both the D-NoB and D-wB groups compared to the reference group (4.5% and 4.1% vs. 0.8%, p<0.001) and length of stay was considerably longer (10.6 and 8.3 days vs. 4.8 days, p<0.001). Individual case characteristics of patients who died in the D-NoB and D-wB groups are presented in Table S2. AKI occurred most frequently in patients with D-wB (26.5%), while patients with D-NoB had AKI less frequently (17.9%) but significantly more than the NoD-NoB group (9.4%, p<0.001). Blood transfusion was received most often in the D-wB group (69.4%) and when transfusion was given, this group received more units on average (4.21 red blood cell units vs. 2.02 in the NoD-NoB group and 2.43 in the D-NoB group) (Table 2). Multivariable logistic regression analysis shows that D-NoB significantly increased the risk of in-hospital death (odds ratio [OR]: 3.45; 95% CI: 1.32, 8.69; p=0.009) while D-wB did not appear significant (OR: 1.25; 95% CI: 0.18, 5.34; p=0.8). Further, transfusion also increased risk of in-hospital death (OR: 10.6; 95% CI: 4.25, 28.2; p<0.001) and baseline anemia was non-contributory in this cohort (Table 3).

**Table 2:** Hospital Outcomes.

| **Variable name** | **NoD-NoB** | **D-wB** | **D-NoB** | **P_Value** | **Overall** |
| --- | --- | --- | --- | --- | --- |
| Transfusion | 8.7% (137/1579) | 69.4% (34/49) | 23.8% (53/223) | <.001 | 12.1% (224/1851) |
| ≥ 2U Transfusion | 47.4% (65/137) | 73.5% (25/34) | 58.5% (31/53) | 0.018 | 54% (121/224) |
| Mean Units Transfusion* | 2.02 +/- 2.11 (137) | 4.21 +/- 3.61 (34) | 2.43 +/- 2.27 (53) | <.001 | 2.45 +/- 2.54 |
| Acute Kidney Injury | 9.4% (149/1578) | 26.5% (13/49) | 17.9% (40/223) | <.001 | 10.9% (202/1850) |
| Length of Stay | 4.84 +/- 5.78 (1579) | 10.59 +/- 7.7 (49) | 8.27 +/- 6.59 (223) | <.001 | 5.41 +/- 6.1 |
| In-Hospital Death | 0.8% (13/1579) | 4.1% (2/49) | 4.5% (10/223) | <.001 | 1.4% (25/1851) |

Values are mean ± SD (n) or %(n).
\* mean transfusion units only among patients who received any transfusion
D-NoB: Hgb drop and no bleed; D-wB: Hgb drop with bleed; NoD-NoB: No Hgb drop and No Bleed (reference group); MI: myocardial infarction; PreProc: pre-procedure

**Table 3:** Multivariable Logistic Regression: In-Hospital Death.

|  | OR | 95% CI | p-value |
| --- | --- | --- | --- |
| **Patient Cohorts** |  |  |  |
| No Hgb Drop-No Bleeding | - | - |  |
| Hgb Drop-Bleeding | 1.25 | 0.18, 5.34 | 0.8 |
| Hgb Drop-No Bleeding | 3.45 | 1.32, 8.69 | 0.009 |
| **Variable** |  |  |  |
| Transfusion | 10.6 | 4.25, 28.2 | <0.001 |
| Anemia | 1.16 | 0.44, 3.29 | 0.80 |

CI: confidence interval; Hgb: hemoglobin, OR: odds ratio

The higher proportion of D-NoB patients having non-femoral or non-percutaneous TAVR approach prompted a sensitivity analysis. Given that blood loss is somewhat inherent to a surgical cutdown approach, bleeding may be inconsistently reported for these procedures; therefore, it is possible that some patients with cutdown were misclassified as D-NoB while in actuality having significant blood loss during their procedures. We performed an analysis of only percutaneous femoral TAVR cases in the cohort (88.6% of total cases). Factors associated with D-NoB were similar to the main analysis (Table S3). Inpatient mortality rate for D-NoB remained higher than the reference group, albeit to a lesser degree, and continued to have a similar hospital mortality compared to D-wB (0.8% vs. 2.4% vs. 2.4%, p<0.001). Figure S1 shows the distribution of subjects in each group over time.

Landmark time-to-event analysis, starting on day of discharge (day 0), demonstrates that mid-term mortality rate was increased in the D-wB patient group (HR: 3.0; 95% CI: 1.63, 5.62; p<0.001), while patients with D-NoB had similar outcomes compared to the reference group (HR: 0.9; 95% CI: 0.55, 1.49; p=0.700) (Figure 1). Multivariate Cox proportional regression analysis shows D-wB maintained a significant association with mid-term mortality (HR: 3.2; 95% CI: 1.83, 5.73; p<0.001) after the addition of clinically relevant variables to the model. Blood transfusion significantly impacted the rate of death on Cox regression analysis (HR: 2.5; 95% CI 1.79, 3.63, p<0.001). AKI was significantly associated with mortality, and increasing BSA was found to be protective (Table 4). Notable non-significant variables included baseline anemia and age. Hgb drop without bleed did not produce an increase in bleeding events at 30-day or 1-year follow-up (log-rank p=0.756) (Figure S2).

**Figure 1.**
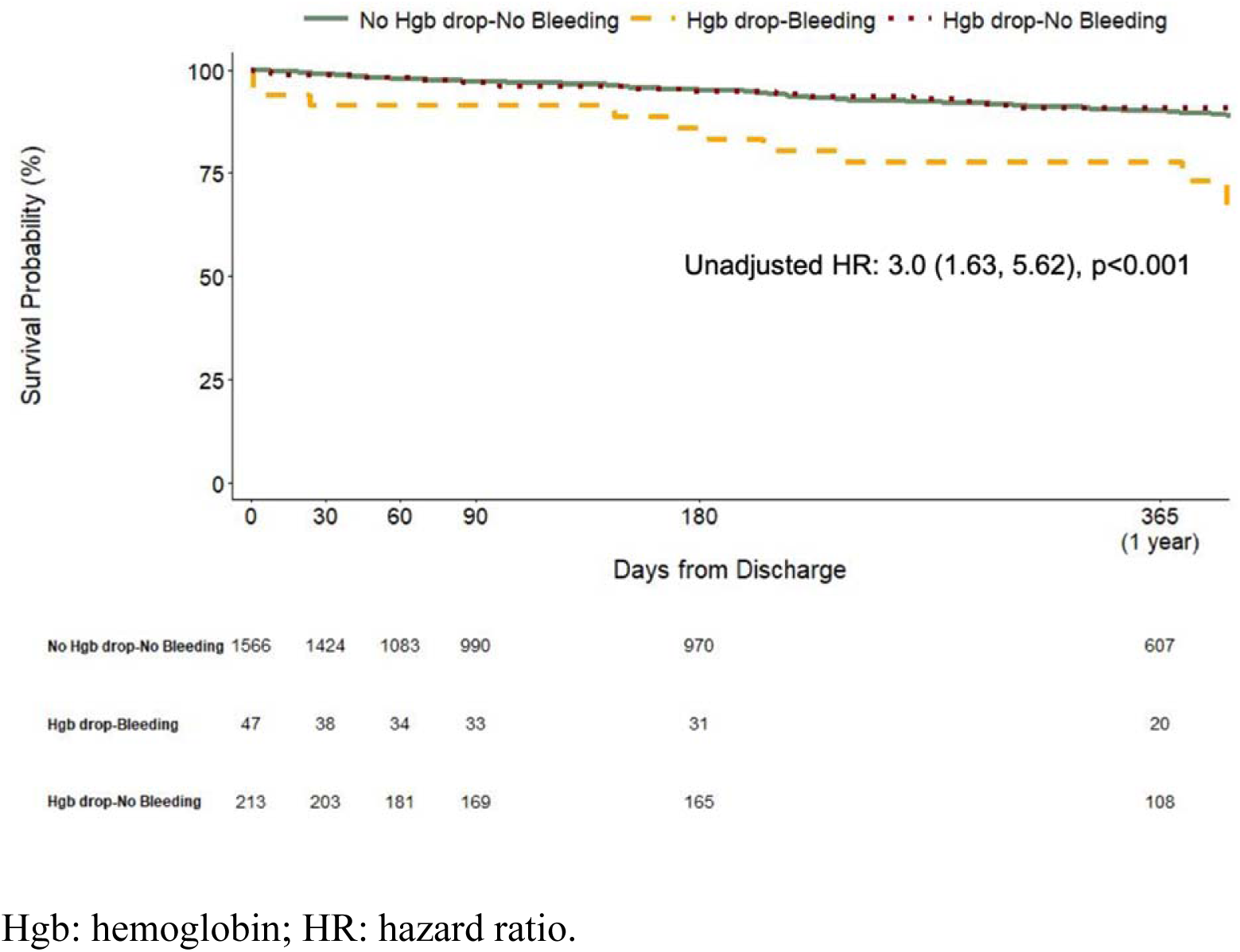
Landmark Kaplan-Meier curve from day of discharge (day 0). Hazard ratio refers to risk of death in the Hgb drop with bleed group compared to the reference group (No Drop-No Bleed cohort).

**Table 4:** Multivariable Cox Proportional Hazard Regression Analysis.

|  | HR | 95% CI | p-value |
| --- | --- | --- | --- |
| **Patient Cohorts** |  |  |  |
| No Hgb Drop-No Bleeding | - | - |  |
| Hgb Drop-Bleeding | 2.25 | 1.15,4.38 | 0.017 |
| Hgb Drop-No Bleeding | 0.83 | 0.49, 1.40 | 0.5 |
| **Variable** |  |  |  |
| Transfusion | 1.52 | 0.99, 2.35 | <0.001 |
| Acute Kidney Injury | 1.78 | 1.16, 2.75 | 0.009 |
| Male Gender | 1.84 | 1.25, 2.72 | 0.0002 |
| Body Surface Area | 0.39 | 0.17, 0.90 | 0.027 |
| Age | 1.02 | 1.00, 1.04 | 0.067 |
| Anemia | 1.42 | 0.95 2.11 | 0.087 |

Hgb: hemoglobin, HR: hazard ratio, CI: confidence interval

## Discussion

This study elucidates the significance of peri-procedural drop in Hgb without bleeding (D-NoB) after TAVR. There were several notable findings: 1) The phenotype of the D-NoB patient is distinct from that of NoD-NoB but similar to patients who have D-wB; D-NoB patients tended towards higher pre-procedure Hgb, smaller body habitus, and advanced age; 2) D-NoB patients were significantly more likely to die peri-procedurally compared to NoD-NoB patients; 3) After hospital discharge, the mortality rate of D-NoB was low, comparable to that of NoD-NoB and diverging from that of D-wB; 4) Blood transfusion, which can be a modifiable risk factor, increased the risk of mortality regardless of bleeding.

TAVR is considered minimally invasive next to its counterpart of surgical aortic valve replacement but still entails a high risk of bleeding due to the need for large-bore femoral access, advancement of the valve along the aorta, and valve deployment under radial force. Each step is a potential source for bleed, which has been shown to profoundly alter patients’ hospital course (re-confirmed by these data)^7^. Within this context, Hgb drop raises particular concern but in many cases does not herald bleeding. Our study found that D-NoB was relatively common, occurring in 12% of cases, 4.6 times the number of patients with D-wB (223 vs. 49).

Hgb drop without bleed is commonly seen among hospitalized patients, though the mechanism is often unclear^4, 8^. In a cohort undergoing TAVR, our data found the D-NoB group to be older, with lower BSA compared to the reference group (NoD-NoB). This finding may suggest that hemodilution is a primary mechanism in D-NoB patients given that degree of hemodilution for a given amount of intravenous fluid administered is dependent on total blood volume, which itself depends on body weight or BSA^9, 10^. Further, pre-procedural Hgb was higher in the D-NoB group, possibly indicating hemoconcentration from dehydration; in this case, intravenous fluids, given routinely or in response to hypotension, would be more likely to stay intravascular, diluting Hgb back to a true baseline. Notably, dehydration is common among elderly or frail patients, especially when kept “nothing per oral” prior to procedure day.

Operators may attempt to avoid the phenomenenon of Hgb drop without bleed by carefully discerning signs of hemoconcentration; these include a change in Hgb level from historic baseline or markers of dehydration prior to the procedure, such as pulse pressure, inferior vena cava collapsibility, laboratory values, or physical exam signs.

The D-NoB group exhibited a higher rate of in-hospital mortality compared to the reference group. Although a decrease in hemoglobin levels without associated bleeding does not directly cause death, it serves as an indicator of heightened periprocedural risk. This is supported by the observed correlation between D-NoB and higher STS-PROM scores, lower BSA, and advanced age, all of which are well-established predictors of peri-procedural mortality. The higher inpatient mortality among D-NoB patients may also be attributed to an increased blood transfusion rate, which could potentially stem from medical teams overreacting to address the drop in hemoglobin levels in the post-TAVR setting. Given that transfusion has been well-documented to correlated with mortality^11–13^, finding supports the notion that blood transfusion should be avoided when possible, i.e., Hgb drop should not immediately trigger transfusion unless high-risk bleeding is suspected (Figure 2). Conversely, sustained anemia in the TAVR population may contribute to bleeding risk and may induce suboptimal cardiometabolics, although anemia in this analysis did not contribute to periprocedural or mid-term mortality. Of note, the transfusion goal in TAVR patients has not been thoroughly defined but appears to favor a restrictive strategy (7-8 g/dL) ^14, 15^.

**Figure 2.**
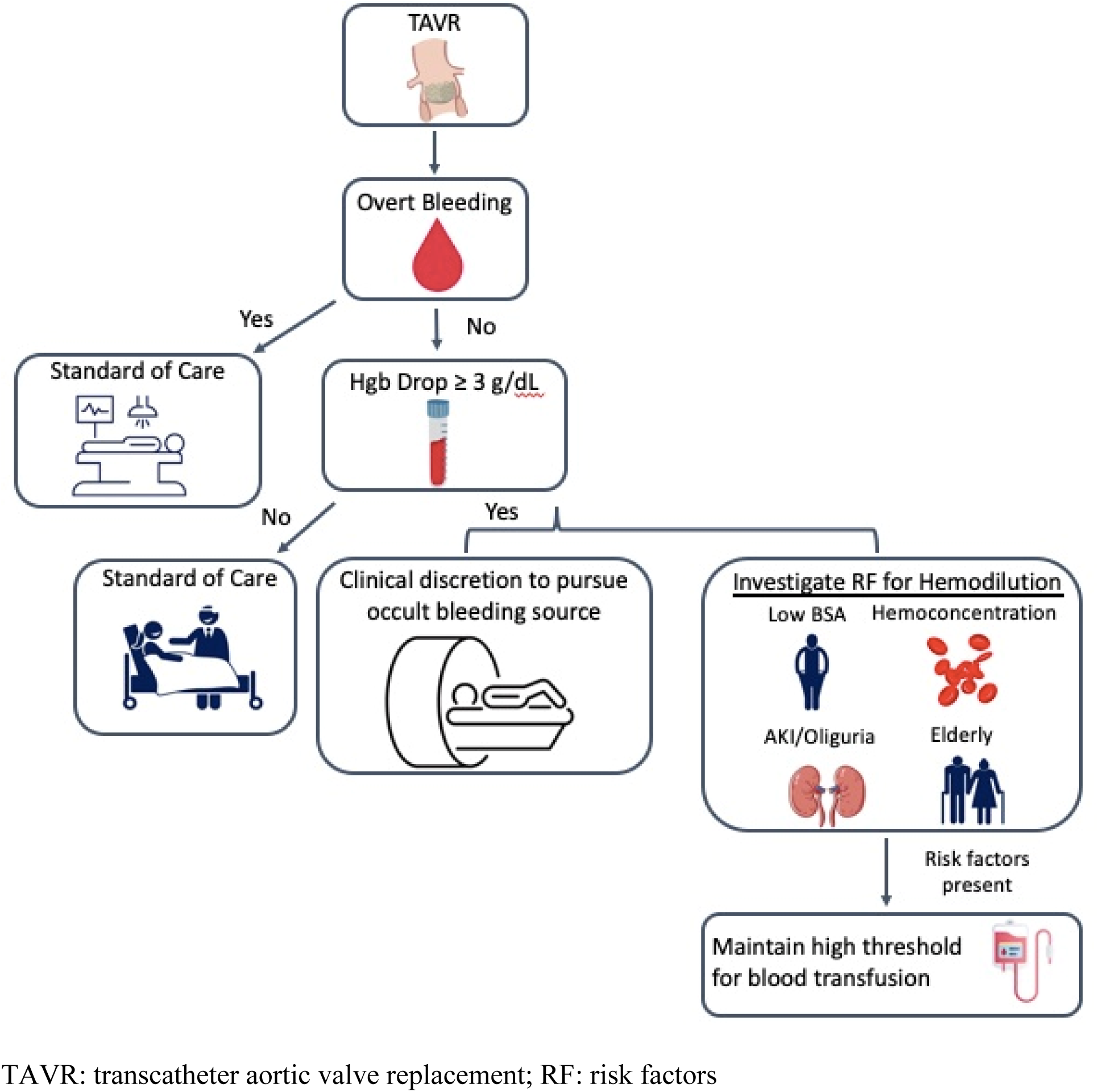
Patient Care Flow Chart for Hemoglobin Drop.

Though in-hospital rate of death was similar between D-NoB and D-wB, mid-term mortality rates among hospital survivors diverged sharply. The difference in mid-term outcomes reinforces the concept that bleeding, while potentially causing acute hemodynamic deterioration leading to short-term death, is also known to initiate a cascade of events that increase the risk of adverse outcomes even after bleeding cessation. These events include ischemia, maleffects of blood transfusion, and de-escalation of antithrombotic regimens^5^.

There are several limitations of this study. First, this was a retrospective analysis for which the usual shortcomings apply. Second, the bleeding event rate was small because our study adhered strictly to the criteria that Hgb drop be at least 3 g/dL. This definition allowed a more precise evaluation of the significance of Hgb drop with vs. without bleeding, but the small event rate did limit statistical analysis of the bleeding group. The apparent lack of effect of bleeding on in-hospital death in multivariable analysis may be attributable to a underpowered sample. Notably, while the DwB sample was small, poor outcomes were in line with established literature. One more limitation is that it is unclear at what time point the patients’ received transfusion during their hospital stay; therefore, the effect of the transfusion on maximum Hgb drop recorded is unknown and the division of groups by Hgb drop from pre-to post-procedure is partially obscured. “Corrected” Hgb was considered; however, without knowing the timing of transfusion, this adjustment would introduce new inaccuracies.

In conclusion, this study confirms that hemoglobin drop without bleeding is common among patients undergoing TAVR. D-NoB represents a patient cohort that is at higher risk of peri-procedural death and, thus, serves as an indicator to provide especially attentive care. Recognizing that advanced age, lower BSA, and hemoconcentration are associated with Hgb drop may allow providers to identify possible dehydration and replete fluids prior to TAVR, potentially avoiding unnecessary testing, AKI, and blood transfusion post-procedure.

### Nonstandard Abbreviations and Acronyms

AKI: acute kidney injury
BSA: body surface area
D-NoB: Hgb drop and no bleed
D-wB: Hgb drop with bleed Hgb: hemoglobin
NoD-NoB: No Hgb drop and No Bleed
TAVR: transcatheter aortic valve replacement

## Data Availability

The data that support the findings of this study are available from the corresponding author upon reasonable request.

## Acknowledgments

This manuscript was last edited by Jason Wermers, MS, a paid medical writer and employee of MedStar Health.

